# Antibiotic Resistance related Mortality, Length of Hospital Stay, and Disability-Adjusted Life Years at select Tertiary Hospitals in Uganda: *A retrospective study*

**DOI:** 10.1101/2024.05.28.24308068

**Authors:** Jonathan Mayito, Flavia Dhikusooka, Daniel Kibombo, Andrew Busuge, Alex Andema, Alfred Yayi, Stephen Obbo, Richard Walwema, Francis Kakooza

## Abstract

**Background:** Antimicrobial Resistance (AMR) is a major global health threat but its burden has not been extensively described in Uganda. We aimed to investigate the antibiotic resistance related mortality, length of hospital stays (LOS) and Disability Adjusted Life Years (DALYs).

**Methods:** This was a retrospective study of clinical records of patients with infectious syndromes admitted at Arua, Jinja and Mbale regional referral hospitals between October 2022 and September 2023. Data was abstracted from clinical records and analyzed to derive the mortality, LOS, DALYs, and factors associated with AMR and mortality using the modified Poisson regression model.

**Results:** Of the 291 participants included in the analysis, 56.6% were males, 10% were living with HIV, and their median age was 35 years (IQR: 21-56). The most common infectious syndromes were sepsis (43.6%) and diarrhea (9.6%). The prevalence of antibiotic-resistant infections (ARI) was 29.6%, while that for multi-drug resistant infections (MDRI) was 17.9%. Patients at Jinja and Mbale RRHs exhibited a 2.42 and 2.65 higher risk of ARI respectively compared to those at Arua RRH. Overall, mortality due to an infectious syndrome was 44.7%, of which 34.9% was due to ARI while 30.8% of the ARI related mortality was due to MDR infections. Most deaths were due to sepsis (42.3%), followed by pneumonia (15.4%) and meningitis (6.2%). Participants with comorbidities had higher risk of mortality due to ARI (45% vs. 31% for those without comorbidities). Among Gram-negative bacteria, *Escherichia coli* and *Klebsiella* spp contributed most to mortality, while among Gram-positives, *Staphylococcus aureus* and *Enterobacter* spp contributed equally. Patients with ARI’s LOS was 1.2 times higher than that for those without ARI while a longer LOS was associated with a 21% higher ARI risk but a 24% decline in risk of death. ARI was associated with higher DALYs, 235.4, compared to 147.0 for non-ARI.

**Conclusion:** The study revealed a high burden of AMR in Uganda, including a high prevalence of MDR and associated mortality, particularly among patients with comorbidities. This indicates an urgent need for stewardship and infection prevention strategies to control the AMR burden.

## Background

The Murry et al 2019 review of the global AMR burden showed a glim picture of accelerated progression towards the 2015 predictions on the impact of antimicrobial resistance (AMR) on global health and economic growth (1). Eight years after the prediction that AMR would kill 10 million people annually by 2050, up to 1.27 million deaths in 2019 were attributed to AMR, surpassing the previous leading infectious killers including tuberculosis, malaria and Human Immuno-deficiency Virus (HIV) (1). Over one million of these deaths occurred in the World Health Organization (WHO) African region, of which majority were in Western Sub-Saharan Africa (2). Worryingly, this burden that disproportionately fell on the Africa region, coincided with poor AMR surveillance systems in the region, where the AMR preparedness score was 53% less than the overall Sub-Saharan Africa 2018-2020 Joint External Evaluation (JEE) score, with many countries lacking National Action Plans (NAP) to address the AMR scourge (3). Antimicrobial resistance has dire consequences including the use of more expensive medicines to treat infections and increased length of hospital stay, which lead to higher healthcare costs and reduced economic productivity [9]. Moreover, resistant infections make undertaking of medical procedures like organ transplants, surgeries and chemotherapy risky (4). To reverse this AMR ascendance and its impact, interventions will need to be based on a wider evidence base of the AMR burden including the clinical outcomes, AMR related economic burden, and disability, which is scarce in the WHO Africa region (1).

Despite the growing global attention on AMR, there are substantial limitations in our understanding of the burden, its distribution and the determinants at the population level (5). Currently, majority of the available AMR data describe resistance profiles or mechanism of resistance (6). While this data is important for surveillance and design of treatment protocols, data on AMR burden including clinical outcomes like mortality, disability, length of stay, and their determinant is critical in informing planning, resource mobilization and allocation, policy formulation, and the design of targeted control strategies.

Individuals who survive AMR have increased Disability Adjusted Life Years (DALYs) including the years of life lost (YLL) and year lived with disability (YLD). DALYs measure the burden of disease: reduction in life expectancy and diminished quality of life due to the disease (7). Consequently, studying the distribution of DALYs in the community helps to know which communities bear the greatest burden of the disease and to focus interventions on these areas [10]. Cassini et al found that DALYs due to AMR (170 per 100 000) were similar to DALYs due to influenza, tuberculosis, and HIV combined (183 per 100,000) and 25% (127 of 501 DALYs per 100 000) of the burden of health-care-associated infections (HAIs) was due to AMR (8). Up to 67·9% (115 of 170) of the total DALYs were accounted for by four antibiotic-resistant bacteria: third-generation cephalosporin-resistant *Escherichia coli*, Methicillin resistant *Staphylococcus aureus* (MRSA), carbapenem-resistant *Pseudomonas aeruginosa*, and third-generation cephalosporin-resistant *Klebsiella pneumoniae* (8). The data indicates a high disease burden posed by AMR.

Determination of the AMR related mortality, length of hospital stay, DALYs and the influencing factors would complement the routine surveillance data that has been accrued during the implementation of the 2018 – 2023 NAP (9–11), and offer a comprehensive description of the AMR burden in Uganda. This study therefore provides a more comprehensive description of the Uganda AMR burden in terms of attributable mortality, DALYs and the driving factors. This information is critical in the campaign to have the Uganda government adopt AMR surveillance in its health prioritization, planning, resource mobilization and allocation.

### Methodology

This was a retrospective study of the AMR related clinical outcomes, length of hospital stay, and associated factors using clinical records of patients with infectious syndromes admitted at Arua, Jinja and Mbale regional referral hospitals (RRH) between October 2022 and September 2023. The records were accessed between 4^th^ and 23^rd^ October 2023. The researchers had access to personal identification information during the data collection but not afterwards. Arua RRH is located in Northwest-Nile region, about 500 km from Uganda’s capital, Kampala while Jinja and Mbale RRHs are located about 100 km and 400 km respectively from Kampala in the Eastern part of Uganda. Arua, Jinja, and Mbale RRHs serve catchment populations of 3.5, 4.5, and 4.6 million people respectively (12–14).

A data abstraction form was designed, pretested and digitalized using Kobo Toolbox, a free and open-source suite of tools for field data collection. Digitalization included building checks in the abstraction tool to ensure quality of the data collected. A team of clinicians were recruited and trained on the protocol, data abstraction tool, and data collection standard operating procedures before data collection. There was a two-tier review of the data by supervisors and a data manager to ensure completeness of the data collected. Abstraction targeted clinical records of patients with infectious syndromes seen at the participating hospital in the 12 months prior to the study who had culture and sensitivity testing done. The study variables abstracted included referral status, prior antibiotic use, admission date, admission diagnosis, sampling date, comorbidities, antimicrobial susceptibility testing (AST) results, antibiotics prescribed after culture, treatment outcome, length of hospital stay, and discharge date or date of death, among others.

### Sample size consideration

Sample size was determined using the University of San Francisco online sample size calculator (15), based on a pooled prevalence of MRSA of 49% and 35% among HIV infected and HIV uninfected respectively (7) and calculated odd ratios (OD) and relative risk (RR) of 1.78 and 1.4 respectively (16). A sample size of 388 records was sufficient to determine the study variables at 80% power and 95% confidence interval (CI).

### Statistical analysis

Statistical analysis was conducted in Stata 18. Continuous variables were reported as median and interquartile range (IQR) and categorical variables as proportions in terms of frequencies and percentages. The association between having or not having an Antibiotic Resistant Infections (ARI) and other categorical variables was determined using the Chi-square test of independence while for continuous variables, we utilized the Two-sample Wilcoxon rank-sum (also known as the Mann-Whitney) test. The resulting p-values provided the level of statistical significance for each variable examined. A modified Poisson regression model was fitted on complete cases (N=224) to determine factors associated with ARI, and Prevalence Ratios at 95% CI were obtained. Factors with p-value < 0.2 in the un-adjusted model were selected into the adjusted model. The DALYs were calculated as a sum of YLL and YLD. YLL was determined as number of deaths multiplied by the standard life expectancy at age of death. Since we dealt with individual data, YLL was taken as the average standard life expectancy at age of death. YLD was determined as the number of new cases of a disease multiplied by Disability Weight (DW) multiplied by the average time a person lives with a disease before remission. The length of hospital stay was obtained as the difference between the date of discharge or date of death and date of admission. At all levels of comparisons, a p-value of < 0.05 was taken as statistically significant.

### Ethical statement

The study was approved by the Makerere University School of Biomedical Sciences Institutional Review Boards (SBS-2023-329), which granted a waiver of consent to use the clinical records of patients with infectious syndromes at the participating hospitals. The study was also approved by the Uganda National Council of Science and Technology (HS3088ES).

## RESULTS

Data was abstracted from 392 records but 101 records for tuberculosis patients were excluded from this analysis because they heavily skewed the data because of the characteristic long length of stay in hospital. In *table 1*, socio-demographic characteristics of 291 participants were examined. Among them, 52.6% were males, 10% were living with HIV, and the median age was 35 years (IQR: 21-56 years).

**Table 1:**
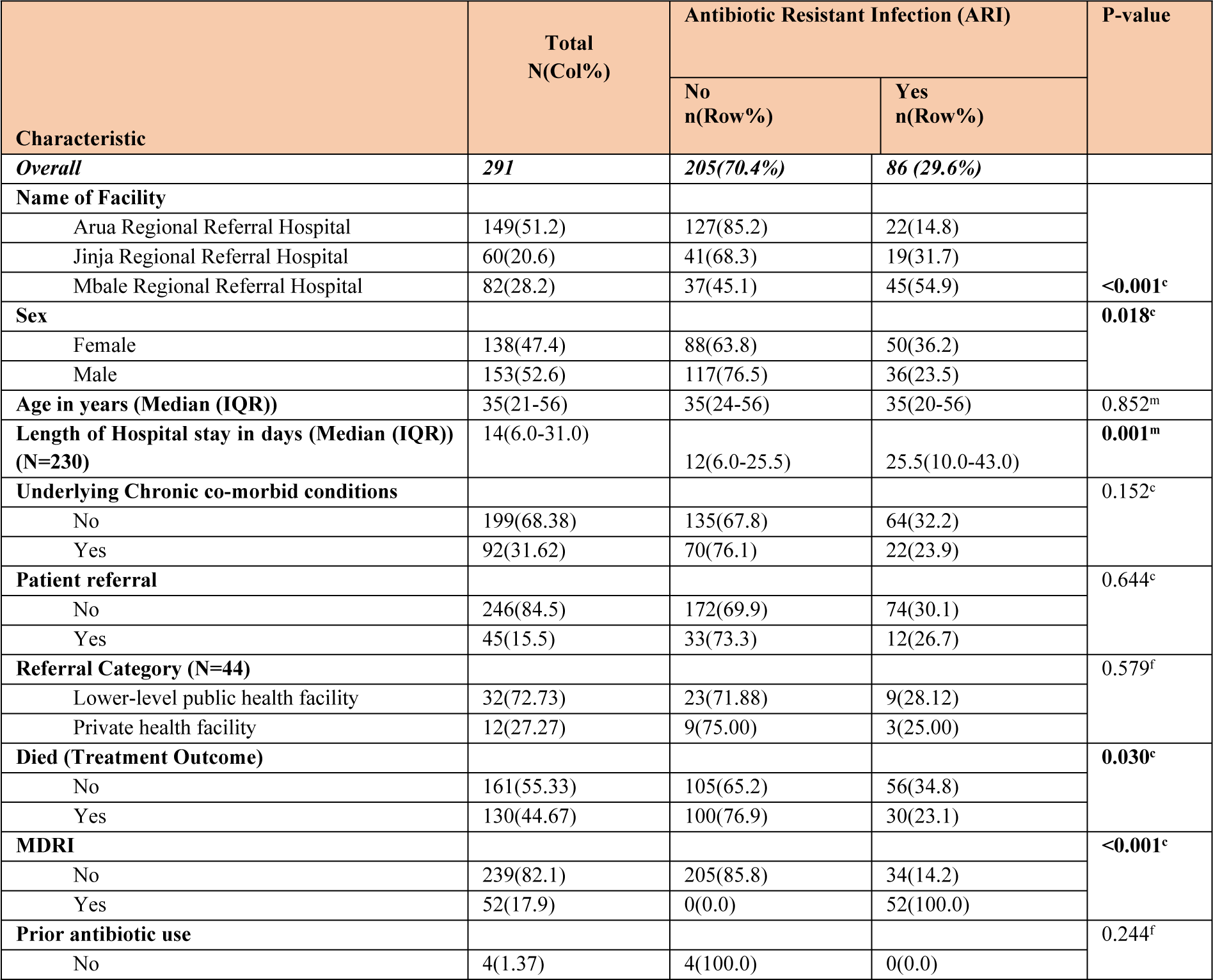
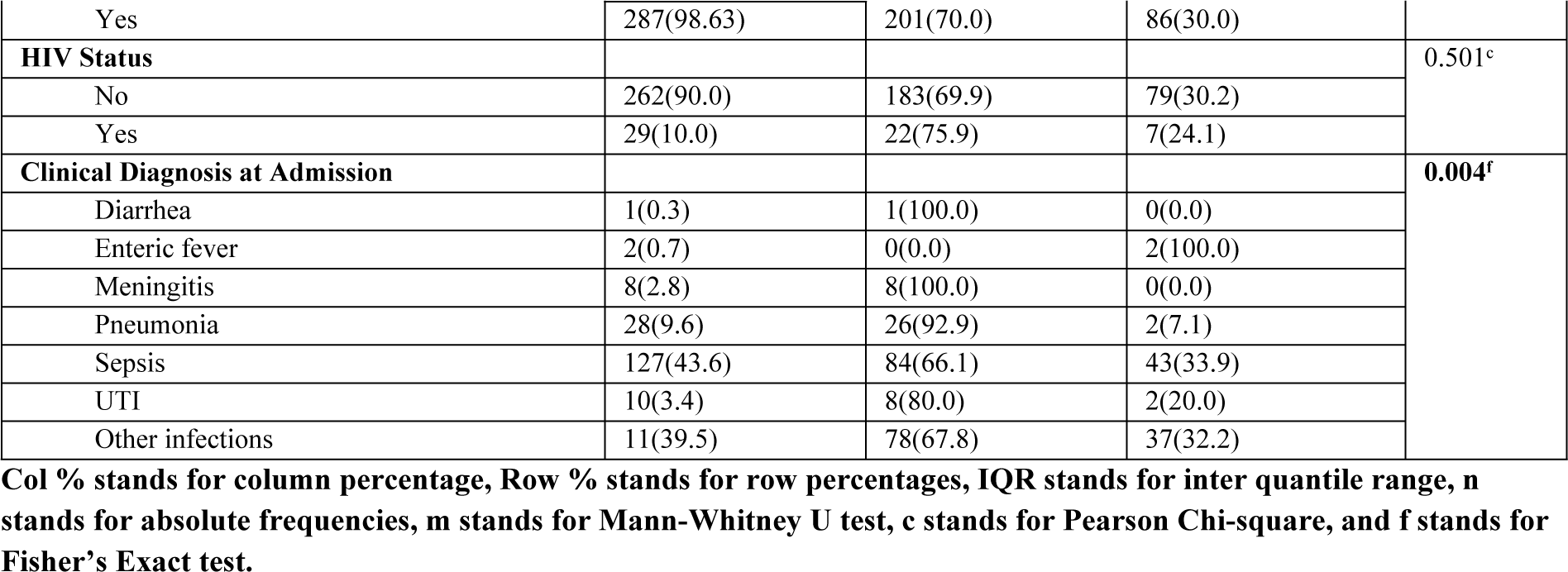
Distributions of demographic characteristics and other factors by antibiotic resistant infections.

### Prevalence of ARI and mortality

The prevalence ARI was 29.6%, while that for multi-drug resistant infections (MDRI) was 17.9%. Overall, mortality due to an infectious syndrome was 44.7%, of which 34.9% was due to ARI while 30.8% of the mortality due to ARI, the causative pathogens were multidrug resistant (MDR), *table 2*. The 30-day mortality was 23% compared to 30% for length of stay longer than 30 days, *table 3*. Additionally, 31.6% of participants had underlying comorbid conditions, with HIV (32%) and diabetes (29%) being the most common. Participants with comorbidities had a high mortality (45%) compared to those without comorbidities (31%), *table 4*. Sepsis (43.6%) and diarrhea (9.6%) were the most common infectious syndromes, *table 5*.

**Table 2:** Mortality due to Antibiotic Resistant Infection.

| Characteristic (N=86) | Frequency (Col %) |
| --- | --- |
| **Multi-Drug Resistant Infections (MDRI)** |  |
| No | 239 (82.10) |
| Yes | 52 (17.90) |
| **Died with ARI (n=86)** |  |
| No | 56 (65.10) |
| Yes | 30 (34.90) |
| **Died with MDRI (n=52)** |  |
| No | 36 (69.20) |
| Yes | 16 (30.80) |

**Table 3:** 30-day mortality due ARI.

|  |  |
| --- | --- |
| **<30 day mortality** |  |
| Number of people who died due to ARI with <30 day LOS and ARI | 13 |
| Total number of people that died wit ARI | 56 |
| % Mortality | 23% |
| **>30 day mortality** |  |
| Number of people who died due to ARI with >30 day LOS and ARI | 17 |
| Total number of people that died wit ARI | 56 |
| % Mortality | 30% |

**Table 4:** Mortality among those with comorbidities.

| **Mortality of patients with comorbidities that also had ARI** | Frequency (Col %) |
| --- | --- |
| Number of patients who had ARI, comorbidities and also died | 10 |
| Number of patients that had ARI and comorbidities | 22 |
| Mortality due to ARI for patients with comorbidities | 45% |
| **Mortality of patients with ARI but had no comorbidities** |  |
| Number of patients who had ARI, and died but had no comorbidities | 20 |
| Number of patients that had ARI and no comorbidities | 64 |
| Mortality due to ARI for patients with comorbidities | 31% |

**Table 5:** Clinical Syndromes contributing to death.

| **Clinical Syndrome** | **Number of Deaths** | **Percentage** |
| --- | --- | --- |
| Sepsis | 55 | 42.3 |
| Other infections | 43 | 33.1 |
| Pneumonia | 20 | 15.4 |
| Meningitis | 8 | 6.2 |
| Enteric fever | 2 | 1.5 |
| UTI | 2 | 1.5 |

### Causes of mortality

Most deaths were due to sepsis (42.3%) followed by pneumonia (15.4%) and meningitis (6.2%). Among the Gram-negative bacteria, *Escherichia coli* (7.8%) followed by *Klebsiella* spp (5.5%) contributed most to mortality while among the Gram-positives, *Staphylococcus aureus* and *Enterobacter spp* contributed equally (2.3%) to mortality. Notably, 85.7% of the deaths were due to ESKAPE organisms (*Staphylococcus aureus*, *Klebsiella spp*, *Pseudomonas aeruginosa*, and *Enterobacter* spp), *table 6*.

**Table 6:**
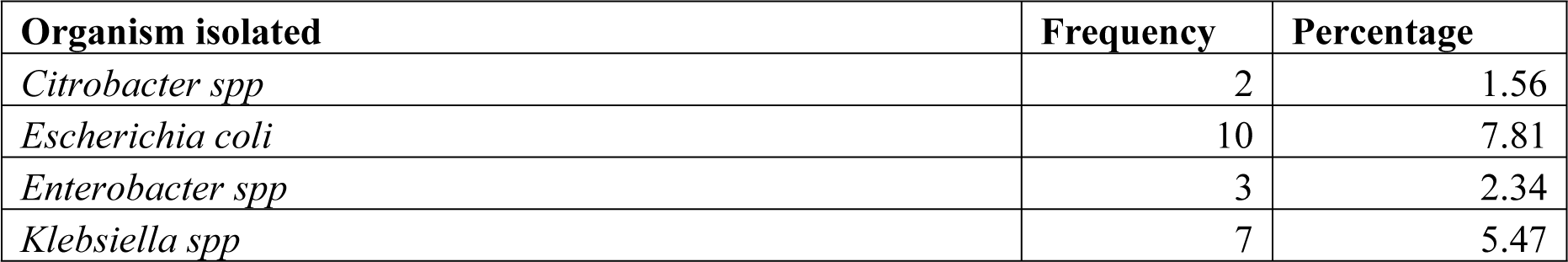
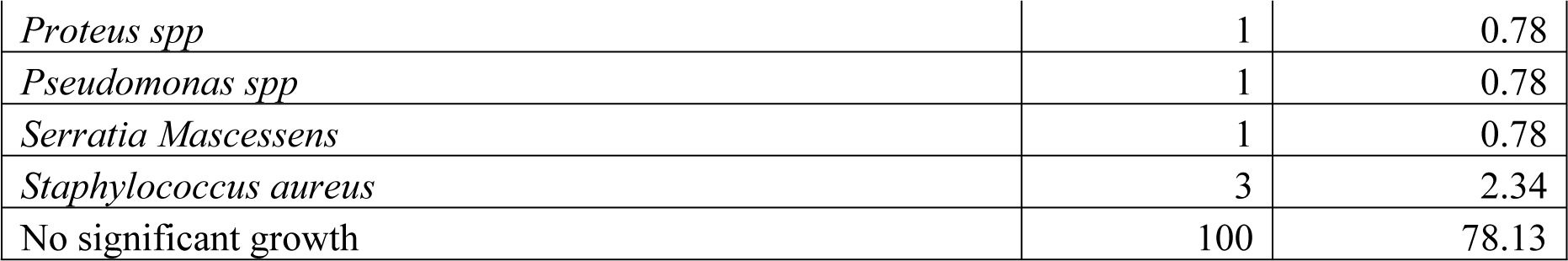
Organisms Contributing to Mortality.

### Factors associated with ARI

Patients at Jinja RRH exhibited a 2.42 times higher likelihood of ARI compared to those at Arua RRH (95% CI: 1.43-4.12, p=0.001). Similarly, patients at Mbale RRH had a 2.67 times higher likelihood of ARI compared to those at Arua RRH (95% CI: 1.53-4.58, p < 0.001). A log unit increase in length of hospital stay in days was associated with 21% higher risk of ARI (95% CI: (1.15-1.95), p=0.003) while mortality was 1.4 (95% CI: 1.00-1.97) times higher among the ARI but this was not statistically significant, *table 7*. For consideration of length of stay and risk of death, ARI related death ratio was 0.76(95% CI: 0.69-0.85), p<0.001 for the log-transformed length of hospital stay, which indicated that for every one-unit log increase in the length of hospital stay, the death rate decreased by approximately 24% (p<0.001, 95% CI: (0.69-0.85)) for patients, *table 7*.

**Table 7:** Factors associated with having an antibiotic resistant infection.

| Characteristic | Un-adjusted PR (95% CI) | P-Value | Adjusted PR (95% CI) | P-value |
| --- | --- | --- | --- | --- |
| **Name of facility** |  |  |  |  |
| Arua Regional Referral Hospital | Reference |  | Reference |  |
| Jinja Regional Referral Hospital | 2.14(1.25-3.67) | **0.005** | 2.42(1.43-4.12) | **0.001** |
| Mbale Regional Referral Hospital | 3.72(2.41-5.73) | **<0.001** | 2.65(1.53-4.58) | **<0.001** |
| **Sex** |  |  |  |  |
| Female | 1.54(1.07-2.21) | 0.019 | 1.49(0.96-2.32) | 0.075 |
| Male | Reference |  | Reference |  |
| **Length of hospital stay (per unit log increase)** | 1.36(1.19-1.55) | **<0.001** | 1.50(1.15-1.95) | **0.003** |
| **Age (Years)** | 1.00(0.99-1.01) | 0.852 | N/A | N/A |
| **Underlying chronic comorbid condition** |  |  |  |  |
| No | Reference |  | Reference |  |
| Yes | 0.74(0.49-1.12) | 0.164 | 0.71(0.44-1.13) | 0.148 |
| **Patient referral** |  |  |  |  |
| No | Reference |  | N/A | N/A |
| Yes | 0.89(0.53-1.49) | 0.651 | N/A | N/A |
| **Died (Treatment outcome)** |  |  |  |  |
| No | Reference |  | Reference |  |
| Yes | 0.66(0.45-0.97) | **0.034** | 1.31(0.88-1.96) | 0.189 |
| **HIV status** |  |  |  |  |
| Negative | Reference |  | N/A | N/A |
| Positive | 0.80(0.41-1.57) | 0.517 | N/A | N/A |

PR denotes Prevalence Ratios and 95% CI denotes the 95% Confidence interval obtained by fitting a modified Poisson regression model on complete cases (N=224). Factors were selected into the adjusted model if $p\text{-value}<0.2$ at un-adjusted.

### Disability adjusted life years (DALYs)

The DALYs for participants with ARI were 235.4, higher than the DALYs for participants without ARI, 147.0.

## Discussion

We provide the first description of bacterial AMR burden including mortality and DALYs in Uganda, a low-income country. Our evaluation reveals a high mortality: 34.9% due to ARI, of which 30.8% was due to MDRI. Moreover, this mortality occurred in presence of a high rate of comorbidities including HIV (32%) and diabetes (29%) and these patients with comorbidities had a higher mortality (45%) compared to those without (31%). Hospital length of stay was 1.2 times higher for patients with ARI compared to those without. A unit increase in length of hospital stay was associated with a 24% reduction in mortality, indicating early mortality before or shortly after AMR has been diagnosed. Most death were due to sepsis, mainly as a result of Gram-negative bacteria, *Escherichia coli* and *Klebsiella spp* in particular as well as *Staphylococcus aureus and Enterococcus* spp among the Gram-positives, with the ESKAPE group accounting for 85.7% of the deaths. All these contributed to higher YLD and DALYs associated with ARI: 189 and 235.4 compared to 101.7 and 147.0 for non-ARI respectively.

We noted a high AMR related mortality, with a significant proportion being due to MDRI, which is similar to what has been observed elsewhere. Globally, AMR is surpassing previously leading infectious causes of death: 1.27 million deaths and 4.95 million deaths were related or associated to AMR in 2019, higher than that due to malaria, tuberculosis or HIV/AIDS (1). In the same year, in the WHO African region, 1.05 million deaths and 250,000 deaths were due to and attributed to AMR respectively, the highest mortality rate (23.5 per 100,000) globally (2). Resistant *Escherichia coli*, among the leading six pathogens causing mortality in the same evaluation, has been associated with an absolute risk of mortality of 58 – 130 per 1000 regardless of the type of resistance (17). South Asia shares with Sub-Saharan Africa the brunt of the AMR burden. Overall, case fatality among neonates admitted to the Neonatal Intensive Care Unit (NICU) in Vietnam was 44.4% with a 31.8% 30-day mortality rate, a 27% increase in odds of fetal outcome and a 2.1 day increase in hospital length of stay (18). A systematic review by Kassim et al reported that AMR increased the mortality due to bloodstream infection by 58% and doubled the risk of ICU admissions (19). The risk of death due to AMR affects both community acquired and hospital acquired (CAI, HAI), where the resistant infections in either type have a higher risk of death (20, 21). AMR has been hitherto an unrecognized infectious cause of death in Uganda. In 2019, up to 7100 deaths were due to AMR while another 30700 deaths were associated with AMR, leaving Uganda with the 165^th^ highest age standardized AMR related mortality among 204 countries (22). Moreover, the AMR related mortality was higher than the previously leading causes of death including tuberculosis, HIV/AIDS and Malaria (22). In this study, the Gram-negative bacteria (*Escherichia coli* and *Klebsiella* spp) were responsible for most of the deaths while *Staphylococcus aureus* and *Enterococcus* spp contributed most among the Gram-positive. These were among the six leading causes of death in 2019 while others included *Streptococcus pneumoniae*, *Acinetobacter baumannii*, and *Pseudomonas aeruginosa* (1). In the African region, *Streptococcus pneumoniae*, *Klebsiella pneumoniae*, *Escherichia coli*, and *Staphylococcus aureus*, were the four leading causes of death while third-generation cephalosporin-resistant *Klebsiella pneumoniae* and methicillin-resistant *Staphylococcus aureus* (MRSA) were the leading pathogen–drug combinations associated with mortality (2). These findings highlight an urgent need to address the AMR challenge, targeting the pathogens responsible for the highest burden in Uganda and the African region at large.

Diabetes and HIV/AIDS were highly prevalent among AMR patients, and although presence of comorbidities was not independently associated with AMR, it carried a heightened risk of death. Hyperglycemia impairs the immunity (23), increasing the risk for infectious syndromes like skin and soft tissue infections, urinary tract infections, surgical site infection, and respiratory tract infections among others (24). Relatedly, drug resistant infections are also increased, for instance multidrug-resistant *Klebsiella pneumoniae* and MRSA were more common in hyperglycemic patients compared to normoglycemic patients (25). There has been contrasting evidence on the risk of AMR posed by diabetes: higher AMR was observed in diabetic patients with liver abscess (26) while carbapenem-resistant *Klebsiella. pneumoniae* was lower among diabetics compared non-diabetics (27). High resistance has been noted in common bacteria isolated from diabetic foot: *Staphylococcus aureus* against gentamicin (57.96%), and ciprofloxacin (52.45%) and *Escherichia coli* and *Klebsiella Pneumoniae* resistance against common antibiotics at more than 50% (28). Overall, AMR is likely to have a higher impact in diabetics due to their heightened risk of infection and frequent exposure to healthcare environment (29). HIV/AIDS was also highly prevalent among patients with AMR in our study. HIV/AIDS weakens the immune system, making individuals prone to infections in addition to the frequent visits to healthcare facilities, which exposes then to infections. HIV/AIDS is associated with high rates of AMR (30): a systematic review showed that people living with HIV (PLWH) were twice as likely to be colonized (31) or infected with MRSA, *Streptococcus pneumoniae* resistant to penicillin, and third-generation cephalosporins resistant *Escherichia coli* and *Klebsiella pneumoniae* (32). Infection prevention measures targeting individuals and hospital or home environment (33), and stewardship practices in the use of antibiotics, in addition to optimized HIV/AIDS care to ensure recovery and sustenance of a robust immune system, are critical measures to protect PLWH from AMR.

On the other hand, we had only few cancer patients in the study as the study was done in a general ward setting. Cancer is a recognized immune suppressive condition including the immune suppressive effect of its treatment while surgery or bone marrow related cancer treatments rely on effective infection prevention or treatment (34). Fifty percent (50%) of cancer deaths are due to infections but deaths due to AMR have not been well described (35). However, high rates of AMR especially in ESKAPE organisms, have been reported in cancer patients: Vancomycin Resistant *Enterococcus* (VRE, 80%), fluoroquinolone resistant *Escherichia coli* (70%), and MRSA (50%), among others (36). Consequences of AMR among cancer patients include increased persistence of bacteremia (25% vs 9.7%), metastatic infection (8% vs 4%), and early case-fatality rates (23% vs 11%) among infections due to resistant ESKAPE pathogens versus other bacteria (36). Moreover, MDR is rampant in cancer patients: 40% in ICU patients including 20% *Escherichia coli* (94.4% ESBL), 12% *Staphylococcus aureus* (90.6% MRSA), 12% *Enterococcus faecium* (18.7% VRE), and 6% *Acinetobacter baumannii* (36). The high burden of comorbidities and associated risk of AMR and the related adverse outcomes highlight the urgent need for strategies to curtail the threat posed by the comorbidities.

Our study’s finding that patients with AMR stayed 1.2 times longer in hospital than patients without AMR, is similar to finding by a study in Ghana, where AMR patients stayed 5 days and 8 days longer than patients with susceptible infections and non-infected patients respectively (19, 37). This is far more than what was found in France, where the excess length of stay due to AMR was only 1.6 days, and 2.9 days in Australia (38, 39). This highlights the difference between the first world and low resource settings. In the first world appropriate diagnostics can aid in early identification of AMR and alternative options of treatment are more available. Long stay in hospital is associated with higher consumption of hospital resources, higher expenditure for the patient, loss of economic productivity and risk of mortality. For instance, in the Ghana study, AMR related costs were USD 1300 higher than those due to susceptible infections, with 30% of the costs being due to loss of productivity through presenteeism and absenteeism (37). Relatedly, the average AMR related cost was less in France (USD 1173) (38) while 40% of AMR related costs in Europe and the United States were also due to loss of productivity (40). A systematic review by Kassim et al estimated a direct cost per AMR case of USD 12,000 in low and middle income countries (LMICs) (19). Hospital acquired infections are more commonly MDR, posing an intense challenge to healthcare. Mauldin et al found that hospital costs and length of hospital stay due to resistant Gram-negative HAI were 29.3% and 23.8% higher than those due to susceptible Gram-negative bacteria (41). On the contrary, however, Wonziak et al did not find any significant difference in length of hospital stay between drug resistant and drug susceptible CAI in Australia (20), as did Barassa et al (42) in Spain. Suzuki et al also found negligible additional length of stay (0.8 days) attributable to resistant *Escherichia coli* and *Klebsiella* spp compared to the prolonged length of stay already imposed by bacteremia due to the same organisms (43). In our study every unit increase in time in hospital was associated with a 24% reduction in mortality. Much as the effect of AMR on length of hospital stay has been widely studied, its effect on or relation with mortality has not been fully evaluated. The finding of reduced mortality with a unit increase in length of stay may imply early mortality due AMR before appropriate diagnosis is made or before appropriate treatment is instituted, implying that those who survive through the early events have higher chances of survival.

By accounting for healthy life years lost due to premature death and years lost living with disabilities, DALYs offer a better estimate of burden of disease (44). We found that the DALYs due ARI (235) were almost two times higher than those due to non-ARI (147). AMR related DALYs in our setting were close but higher than DALYs observed in Japan (195) in 2021 (45) but two and four times higher than those in Switzerland (98) and German (57) respectively in 2023 (46), highlighting that low resource settings bear a significantly higher AMR burden. Moreover, the magnitude of DALYs can be affected by type of pathogen (*Staphylococcus aureus, Acinetobacter baumannii,* and *Streptococcus pneumoniae* are associated with higher DALYS), sex (higher in male except in women in the reproductive age groups), and site of infection (blood stream infections and bone and joint infection are associated with higher DALYs) while geographical and socio-demographic differences have also been noted (45, 47, 48). These differences likely result from the effect of these parameters on the components used to calculate YLL and YLD particularly number of deaths, incidence of AMR cases, and disability weights. Availability of diagnostic tests and alternative options of treatment for AMR which affect risk of death or the time one stays with the resistant infection, might also explain the difference between high versus low income settings.

We note that the admitting hospital and length of stay in hospital were independently associated with ARI. The three admitting hospitals are located within cities, however, the communities in eastern part of the country are more urbanized than those in the north. Bacterial activity, antibiotic resistant genes, and MDR bacteria in the environment have been noted to increase with urbanization and to differ by level of urbanization, with population size as a key driving factor (49). Conditions in urban centers including; poor housing, poor drainage and poor sanitation; poorly-regulated and private provision of medical services including access to antimicrobials, poor waste management, and increased demand for food-animals associated with inappropriate use of antibiotics as growth-promoters, which drive emergence and transmission drug resistant infection, increase with urbanization (5). Mauldin et al also showed that parameters associated with increased hospital costs and length of stay respectively included pneumonia diagnosis (43.8% vs. 38.2%), ICU admission (142% vs. 106%), neutropenia (83.5% vs. 70.9%) and organ transplant (115.8% vs. 74%) while patients 12 years or more had a 26.3% lower hospital cost but a 66.8% lower length of stay versus those younger (41). Among patients referred for long term care in Chicago, USA, presence of a feeding tube and polymicrobial infection were associated with ARI while urinary catheter, decubitus ulcer, prior use of antibiotics and previous admission within 30 days of current admission were not (50). On the other hand, MDR *Pseudomonas aeruginosa* infection was independently association with sex, age, and comorbidities, and correlated with ventilation history and neutropenia (51). On the other hand, cotrimoxazole, amikacin, or imipenem except cefazolin; admission to ICU and prior use of acid-suppressive medication were associated with infection with MDR organisms (52). These factors can be targeted by strategies to control AMR and those to improve patient outcomes.

Our study strength lies in it being the first expanded description of AMR burden in Uganda, a resource limited setting, revealing critical data that can inform strategies to control AMR, improve clinical outcomes as well as planning and resource allocation. It was limited in geographical scope as only two regions of Uganda were represented, which may not be representative of other regions in the country. Finally, because of the retrospective design, there was missing data on some of the critical parameters that would have been of interest to evaluate.

### Conclusion/recommendation

Uganda faces a high AMR burden characterized by a high mortality and a high rate of MDR infections, high DALYs and increased length of hospital stay, in a setting of a high rate of comorbidities including HIV and diabetes. Sepsis, majorly due to resistant Gram-negative bacteria, contributed most to the AMR burden. While these findings can shape the early targeted AMR control strategies, a larger evaluation covering all regions of the country to comprehensively describe the burden is urgently needed.

## Author contribution

Jonathan Mayito, Conceptualization, Investigation, Methodology, Project Administration, Writing – Original Draft Preparation Flavia Dhikusooka, statistical analysis, Review and Editing Daniel Kibombo, Investigation, Methodology, Writing – Review & Editing Alex Andema, Writing – Review & Editing Stephen Obbo, Writing – Review & Editing Alfred Yayi, Writing – Review & Editing Richard Walwema, Resource, Methodology, Supervision, Writing – Review & Editing Francis Kakooza, Resources, Methodology, Supervision, Writing – Review & Editing

## Conflict of interest

All authors report no conflict of interest.

## Financial disclosure

This work was funded by the Royal Society of Tropical Medicine and Hygiene (RSTMH) Early Career Award in partnership with the National Institute for Health and Care Research (NIHR).

## Data Availability

All data will be availed after acceptance

## Notes

### Competing Interest Statement

The authors have declared no competing interest.

